# Genotype and Clinical Characteristics of Patients with Wolfram Syndrome and WFS1-related Disorders

**DOI:** 10.1101/2023.02.15.23284904

**Authors:** Evan M. Lee, Megha Verma, Nila Palaniappan, Emiko M. Pope, Sammie Lee, Lindsey Blacher, Pooja Neerumalla, William An, Toko Campbell, Cris Brown, Stacy Hurst, Bess Marshall, Tamara Hershey, Virginia Nunes, Miguel López de Heredia, Fumihiko Urano

## Abstract

**Objective:** Wolfram syndrome (WFS) is an autosomal recessive disorder associated with juvenile-onset diabetes mellitus, optic atrophy, diabetes insipidus, and sensorineural hearing loss. We sought to elucidate the relationship between genotypic and phenotypic presentations of Wolfram syndrome which would assist clinicians in classifying the severity and prognosis of Wolfram syndrome more accurately.

**Approach:** Patient data from the Washington University International Registry and Clinical Study for Wolfram Syndrome and patient case reports were analyzed to select for patients with two recessive mutations in the WFS1 gene. Mutations were classified as being either nonsense/frameshift variants or missense/in-frame insertion/deletion variants and statistical analysis was performed using unpaired and paired t-tests and one- and two-way ANOVA with Tukey’s or Dunnett’s tests.

**Results:** A greater number of genotype variants correlated with earlier onset and a more severe presentation of Wolfram syndrome. Secondly, non-sense and frameshift variants had more severe phenotypic presentations than missense variants, as evidenced by optic atrophy emerging significantly earlier in patients with 2 nonsense/frameshift alleles compared with 0 missense transmembrane variants. In addition, the number of transmembrane in-frame variants demonstrated a statistically significant dose-effect on age of onset of diabetes mellitus and optic atrophy.

**Summary / Conclusions:** The results contribute to our current understanding of the genotype-phenotype relationship of Wolfram syndrome, suggesting that alterations in coding sequences result in significant changes in the presentation and severity of Wolfram. The impact of these findings is significant, as the results will aid clinicians in predicting more accurate prognoses and pave the way for personalized treatments for Wolfram syndrome.

## 1 Introduction

Wolfram syndrome (WFS) is an autosomal recessive disorder associated with juvenile-onset diabetes mellitus, optic atrophy, diabetes insipidus, and sensorineural hearing loss [1]. Diagnosis of Wolfram syndrome is usually ascertained due to the occurrence of early onset type 1 diabetes mellitus with optic atrophy, which occur in the first decade of life [1; 2]. Additionally, central diabetes insipidus and sensorineural deafness occur in the second decade, dilated renal outflow tracts occur in the third decade, and neurological symptoms appear in the fourth decade [1]. Patients can also develop a wide range of symptoms including, bladder and bowel dysfunction, temperature regulation defects, gait ataxia, balance deficits, and loss of sense of taste and smell [1; 3; 4]. WFS symptoms have a detrimental impact on patients’ quality of life and daily functioning [3; 4]. The life span of patients is expected to be around 30–40 years of age due to respiratory failure caused by brainstem atrophy [1; 2]. The prevalence of the disorder is estimated to be present in 1 in 100,000 in North America and 1 in 770,000 in the UK [1; 5].

Two causative genes, *WFS1* and *CISD* genes have been implicated in the development of Wolfram syndrome. *WFS1* encodes for wolframin, an endoplasmic reticulum (ER) membrane glycoprotein, which plays a role in Ca2+ homeostasis and regulates the ER stress response [6]. Mutations in wolframin lead to ER and mitochondrial dysfunction which cause apoptosis and cell death [3]. *CISD* encodes for an ER intermembrane small protein (ERIS) which plays a role in Ca2+ homeostasis and mitochondrial function [3; 7]. Mutations in *CISD* were initially described in Jordanian patients with unique phenotypic presentations such as bleeding tendency, defective platelet aggregation with collagen, and peptic ulcer disease [8].

Additionally, pathogenic variants in *WFS1* can cause the development of *WFS1*-related disorders involving sensorineural low frequency hearing loss, hearing loss and optic atrophy, cataracts, and an autosomal dominant syndrome characterized by neonatal diabetes, congenital cataracts, sensorineural deafness, hypotonia, intellectual disability, and development delay [9]. Dominant WFS1 variants potentiate ER stress and result in pathophysiology that is distinct and less severe than patients with recessive Wolfram syndrome [9].

The autosomal recessive syndrome has been characterized widely in literature and a variety of pathogenic variants and polymorphisms have been reported to date. In the *WFS1* variants present in the literature, alterations in coding sequences have identified changes including deletions, insertions, nonsense and missense mutations [10]. Associations between genotype and phenotype characteristics can suggest the role that gene alterations play in the variability of clinical phenotypes. We sought to elucidate the relationship between genotypic and phenotypic presentations of Wolfram syndrome. Additional information about genotype and phenotype correlations would allow clinicians to classify the severity of Wolfram syndrome more accurately. This could aid in predicting more accurate prognoses and pave the way for personalized treatments for Wolfram syndrome.

The advantages to discovering genotype and phenotype correlations are highlighted in the case of another autosomal recessive disorder, cystic fibrosis. Typing the genotype-phenotype relationship for cystic fibrosis is important as pathogenic variants can alter the expression and function of CFTR via multiple mechanisms [11; 12; 13]. Additionally, the diverse clinical consequences of cystic fibrosis can be attributed to modifier genes and the environment in combination with pathogenic variants [11; 14; 15]. As such, accurate classification of *CFTR* variants, as well as causative Wolfram variants, is essential in optimizing the treatment of individuals [11].

In this study, we aim to classify the range of severity of clinical presentations of autosomal recessive Wolfram syndrome. We classify genetic variants by age of onset, type of genetic variant, and location of variant to identify associations with disease severity. Due to the rare prevalence of Wolfram syndrome, there is fragmented data regarding the correlation between genotype and phenotype presentations. To address this, we compiled patient data from the Washington University International Wolfram Syndrome and *WFS1* Related Disorders Registry and patient data from published case reports compiled in a systematic review by Heredia et al [2]. We performed meta-analysis on these data and found significant correlations between pathogenic variant characteristics and disease severity.

## 2 Materials and Methods

### 2.1 Patients

Subjects, and their parents or legal guardians, as appropriate, provided written, informed consent before participating in this study, which was approved by the Human Research Protection Office at Washington University School of Medicine in St. Louis, MO (IRB ID #201107067).

Patient data from the Washington University International Registry and Clinical Study for Wolfram Syndrome and patient case reports highlighted in Heredia et al were analyzed to select for patients with two recessive variants in the *WFS1* gene [2]. Patients were excluded if they lacked genetic information for either of their *WFS1* allele variants. Additionally, records were excluded if they did not have a numerical age of onset for their respective clinical phenotype (diabetes insipidus, optic atrophy, diabetes insipidus, hearing loss). Pathogenic variants were then classified as being either nonsense/frameshift variants or missense/in-frame insertion and deletion variants. The average age of onset was calculated for each clinical phenotype based on the number of nonsense/frameshift alleles. For patients with either 0 or 1 nonsense/frameshift variant, their other variant was classified as transmembrane or not based on whether the amino acid position was in one of the transmembrane domains provided on UniProt, and age of onset was noted.

### 2.2 Statistical Analysis

Statistical analysis was performed by unpaired and paired t tests and one- and two-way ANOVA with Tukey’s or Dunnett’s tests. Statistical tests are specified in figure legends. Error bars on all graphs represent a 95% CI. P < 0.05 was considered statistically significant. Data are shown as means ± SEM unless otherwise noted.

## 3 Results

### 3.1 Onset Age of Clinical Manifestations of Wolfram Syndrome

Average age of onset of clinical manifestations for the combined patient cohort was calculated for each of diabetes mellitus, optic atrophy, diabetes insipidus, and hearing loss (Figure 1). The mean age of onset was 7.5 years (95% Confidence Interval 6.8-8.1 years) for diabetes mellitus, followed by 12.9 years (12.0-13.8 years) for optic atrophy, 14.1 years (12.9-15.3 years) for diabetes insipidus, and 15.3 years (14.0-16.5 years) for hearing loss. The ages of onset are similar to but slightly lower than those in a similar cohort study of 67 Japanese patients with Wolfram Syndrome, which found median age of onsets to be 8.7 years, 15.8 years, 17.2 years, and 16.4 years for diabetes mellitus, optic atrophy, diabetes insipidus, and hearing loss respectively [25].

**Figure 1.**
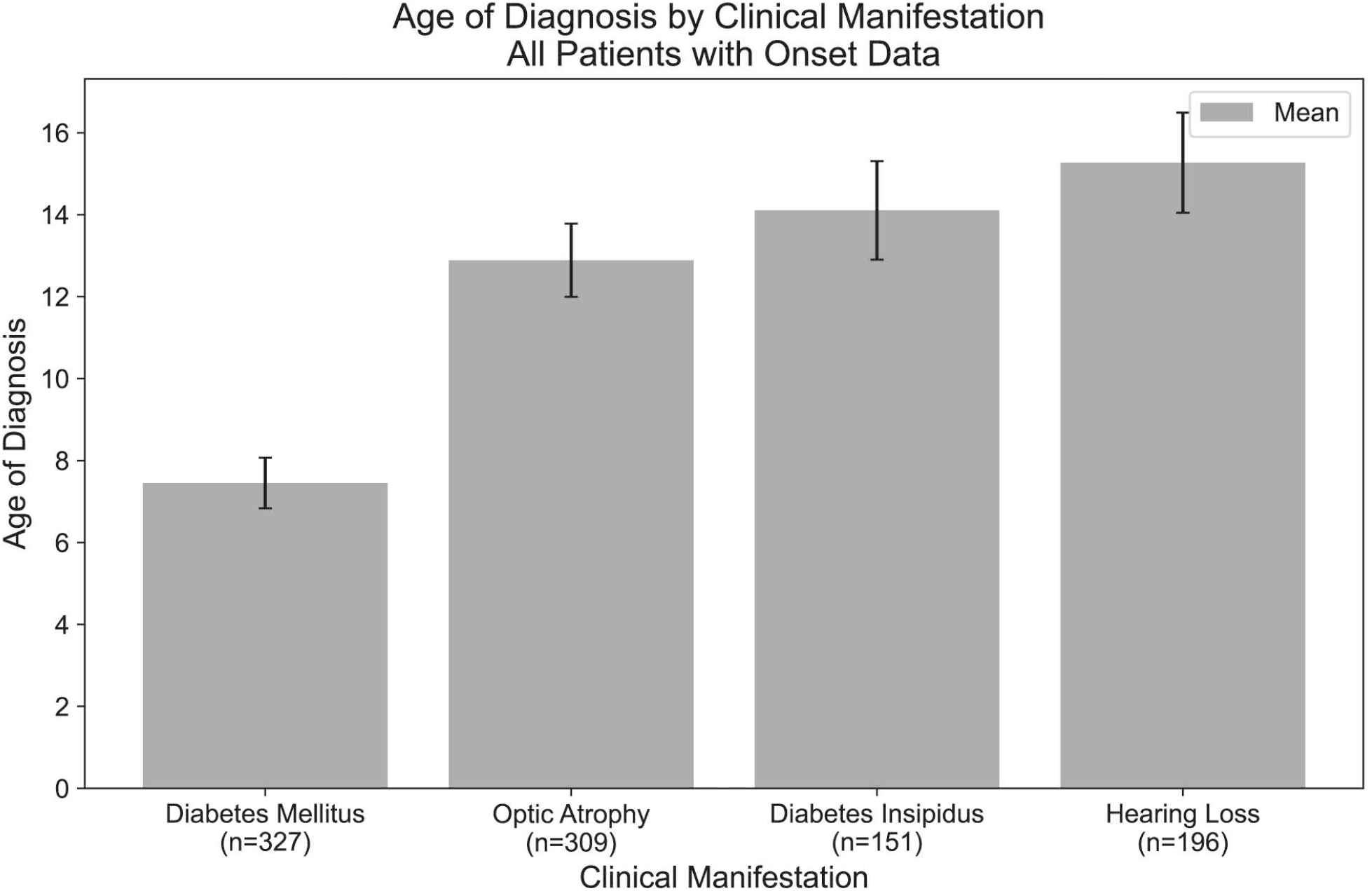
Mean age of onset of clinical manifestations of Wolfram Syndrome. Diabetes mellitus, optic atrophy, diabetes insipidus, and hearing loss emerged respectively at mean ages of 7.5, 12.9, 14.1, and 15.3 years.

### 3.2 Nonsense/Frameshift *WFS1* Alleles Exhibit a Dose-Dependent Response on Disease Severity

For each clinical manifestation of Wolfram Syndrome, patients who had information on both alleles as well as numerical age of onset data were further classified based on whether they had zero, one, or two nonsense/frameshift (NSFS) variant alleles. While diabetes insipidus and hearing loss showed no association of onset age with the number of NSFS variants, both diabetes mellitus and optic atrophy demonstrated a dose-effect of number of NSFS variants with respect to age of onset (Figure 2A). Both diabetes mellitus and optic atrophy emerged earliest in patients with 2 NSFS alleles, followed by patients with 1 NSFS allele, followed by patients with 0 NSFS alleles and only in-frame variants. Diabetes mellitus emerged significantly earlier in patients with 2 NSFS alleles compared with both 0 and 1 NSFS alleles, and correspondingly emerged at a mean age of 5.3 years, 7.7 years, and 9.8 years for 2, 1, and 0 NSFS alleles. Optic atrophy emerged significantly earlier in patients with 2 NSFS alleles compared with both 0 and 1 NSFS alleles, and correspondingly emerged at a mean age of 10.6 years, 13.7 years, and 15.1 years for 2, 1, and 0 NSFS alleles. One outlier to this finding is emphasized in an exceptional case of typical wolfram syndrome. This patient has a phenotypical clinical diagnosis of typical Wolfram Syndrome with the development of diabetes mellitus at age 5 and optic atrophy at age 8. However, we could only detect a frameshift pathogenic variant in one allele with a corresponding normal allele. Since we could not detect two pathogenic variant alleles, we hypothesized the normal allele was not expressed. This prompted us to look at the expression levels of wild-type and mutated alleles by next-generation sequencing using RNA extracted from induced pluripotent stem cells (iPSC) derived from this patient. Results showed no expression of the wild-type allele, suggesting that there is suppression or methylation in the gene regulatory region of the normal allele. There was not a statistically significant difference in the age of onset between patients with 0 NSFS alleles and 1 NSFS allele for either diabetes mellitus or optic atrophy. The mean onset, confidence intervals, number of patients in each category, and multiple-test-corrected q-values are shown in Table 1.

**Figure 2.**
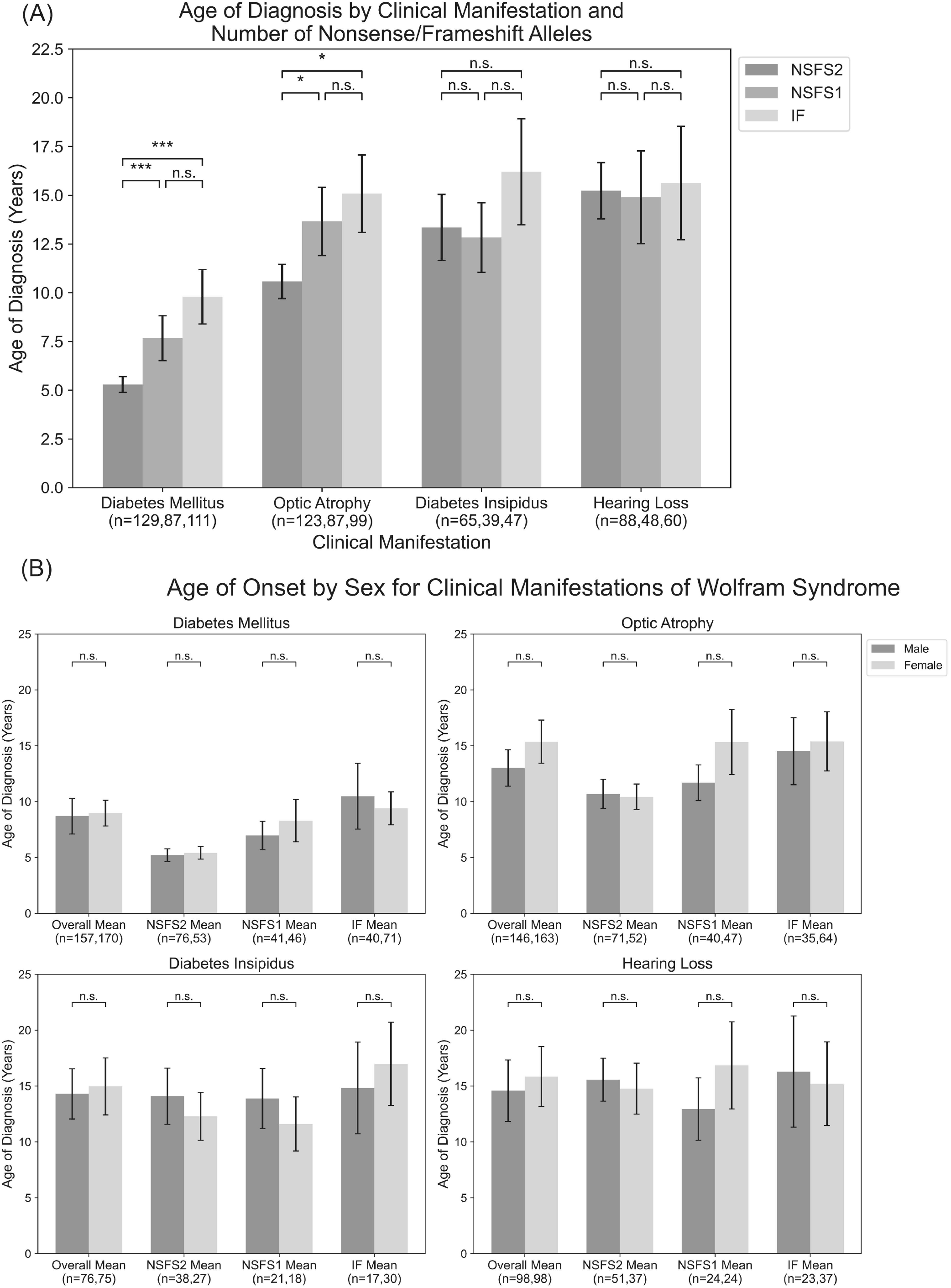
Analysis of genotype-phenotype correlations for number of nonsense/frameshift variants. Age of onset of different clinical manifestations by number of nonsense/frameshift variants vs in-frame missense or insertion/deletion variants for the entire dataset (A) and split by patient sex (B). NSFS2: two nonsense/frameshift variants (n=65-129). NSFS1: one nonsense/frameshift variant (n=39-87). IF: no nonsense/frameshift variants (n=47-111). *, *P*<0.05; **, *P*<0.01; ***, P<0.001; n.s., no significance. P-values were assigned using Wilcoxon’s Rank Sum Test with multiple test correction applied via the Bonferroni method.

No statistically significant differences were observed in age of onset between male and female patients, either within the above number of NSFS variant categories or for the cohort overall (Figure 2B).

### 3.3 Missense *WFS1* Variants in Transmembrane Domains Are Associated with Earlier Onset of Disease

For missense and in-frame insertion and deletion variants, amino acid position information was matched against the predicted transmembrane domain annotations from UniProt. These variants were then clarified as transmembrane (TM) or non-transmembrane (NTM). For each clinical manifestation, the subset of patients previously defined with a) 0 NSFS alleles and consequently two in-frame variants and b) 1 NSFS allele and 1 in-frame variant were then further classified by whether their in-frame variant was TM or NTM. Average age of onset for each clinical manifestation was compared by the number of TM alleles (Figure 3A). Patients with two in-frame variants (0 NSFS variants) showed a similar effect of the number of transmembrane variants as seen with the number of NSFS alleles, with the number of in-frame variants which were transmembrane demonstrating a statistically significant dose-effect on age of onset of DM and OA. Diabetes mellitus emerged significantly earlier in patients with 2 TM variants in 2 missense/in-frame variants compared with both 0 and 1 NSFS variants, and correspondingly emerged at a mean age of 6.3 years, 9.0 years, and 13.5 years for 2, 1, and 0 TM variants. Optic atrophy emerged significantly earlier in patients with 2 NSFS alleles compared with patients that had 2 missense/in-frame variants with 0 TM variants. There was no statistically significant difference in the age of onset between patients with 2 NSFS alleles and 1 TM variant for optic atrophy. Optic atrophy correspondingly emerged at a mean age of 10.7 years, 14.4 years, and 19.3 years for 2, 1, and 0 NSFS alleles. The mean onset, confidence intervals, number of patients in each category, and p values are shown in Table 2.

**Figure 3.**
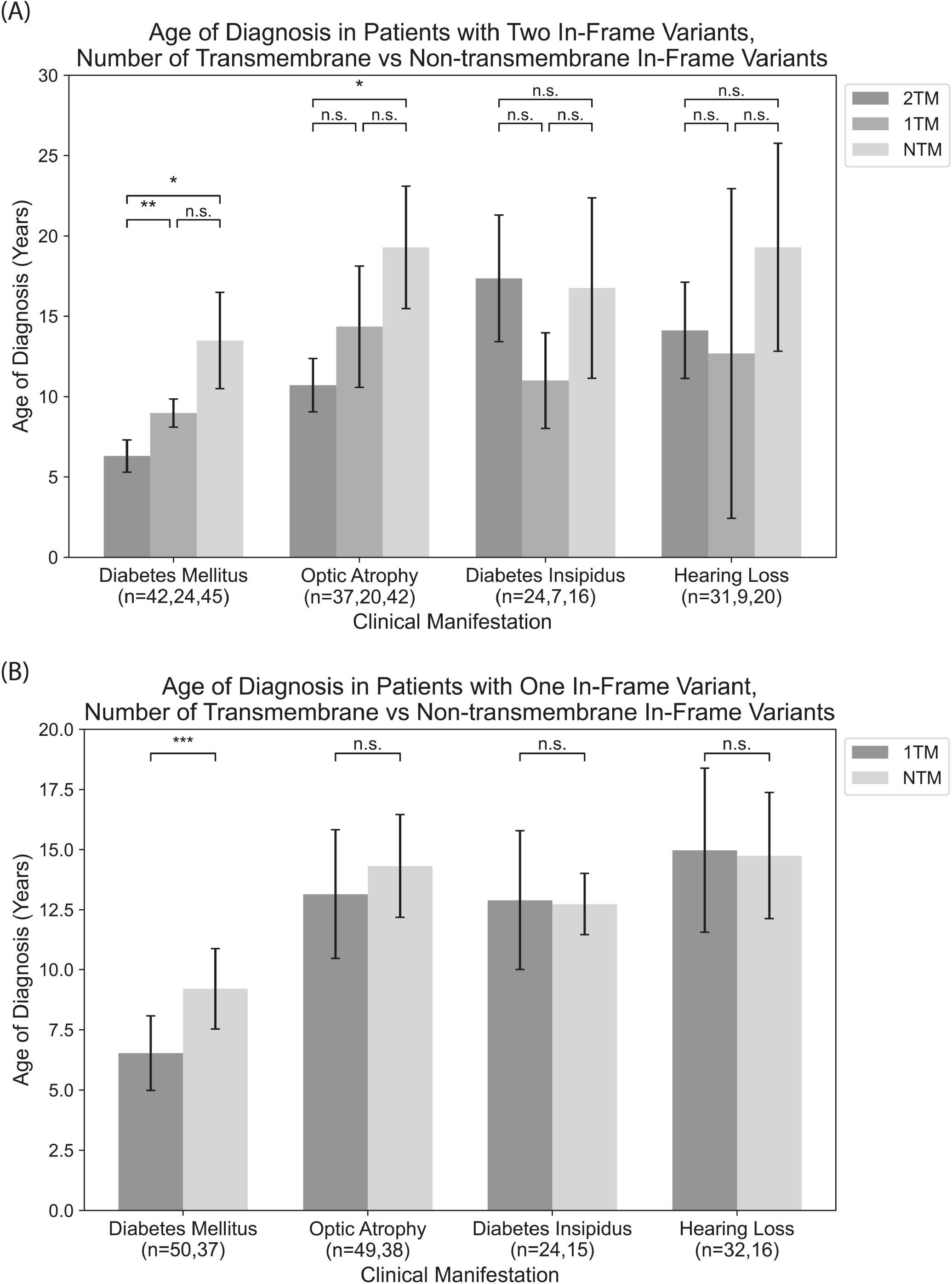
Analysis of genotype-phenotype correlations for in-frame variants by transmembrane/non-transmembrane domain. Age of onset of different clinical manifestations by number of in-frame variants (either missense or in-frame insertion/deletion) in transmembrane domains for patients with two in-frame variants (A) and patients with one in-frame variant (B). 2TM: Two transmembrane variants (n=24-42). 1TM: One transmembrane variant (n=7-24). NTM: No transmembrane variants (n=16-45). *, *P*<0.05; **, *P*<0.01; n.s., no significance. P-values were assigned using Wilcoxon’s Rank Sum Test with multiple test correction applied via the Bonferroni method.

In patients with one in-frame variant and one nonsense/frameshift variant, an in-frame variant in a TM position had statistically significant earlier onset of diabetes mellitus but none of the other clinical manifestations (Figure 3B).

## 4 Discussion

In the setting of Wolfram syndrome, we sought to explore the associations between genotype and phenotype characteristics to speculate the role that gene alterations can play in the variability of clinical phenotypes. It was found that both diabetes mellitus and optic atrophy demonstrated a dose-effect of number of NSFS variants with respect to age of onset. In addition, the number of transmembrane in-frame variants demonstrated a statistically significant dose-effect on age of onset of DM and OA.

These results highlight principles that are important in understanding the genotype-phenotype relationship in Wolfram syndrome. Firstly, it is suggested that a greater number of variants correlates with earlier onset and a more severe presentation of Wolfram. The relationship between variant characteristics and disease severity has been explored previously with the discovery of the Arg558Cys mutation in Ashkenazi Jew individuals which is associated with a milder, late-onset phenotype of Wolfram syndrome including early onset diabetes and reduced penetrance for optic atrophy [16; 17].

Secondly, it is suggested that non-sense and frameshift mutations have more severe phenotypic presentations than missense mutations, as evidenced by optic atrophy emerging significantly earlier in patients with 2 NSFS alleles compared with 0 missense TM variants. A similar phenomenon is highlighted in the case of another autosomal recessive disorder, cystic fibrosis. Literature on genotype-phenotype correlations in cystic fibrosis predicts milder phenotypes and better prognosis associated with A455E, a missense mutation, compared to ΔF508, a deletion mutation [18].

Additionally, it is suggested that if missense mutations are present in the transmembrane domain, there is predicted to be an earlier age of onset for the traits of DM and OA. Possible explanations for this include missense mutations in the TM domain making protein misfolding and aggregation more likely. Pelizaeus–Merzbacher disease is an early-onset leukodystrophy caused by frameshift and missense mutations of the PLP1 gene. Mutations can lead to protein misfolding and retention of PLP and DM20 (proteins in the TM domain) in the ER, supporting the conjecture that Wolfram mutations in the TM domain contribute to protein misfolding and aggregation [19].

Lastly, there are no differences observed in the age of onset of DM and OA between male and female patients. This finding is surprising as the pathophysiology of Wolfram syndrome involves ER stress and estrogen has been shown to mitigate ER stress [20]. Additionally, in *WFS1* knockout mice, males have been shown to present with more severe phenotypes than females [21]. These findings raise the possibility that female patients would present with milder manifestations of Wolfram syndrome; however, this hypothesis was not supported by our data.

The impact of these findings is significant as the results offer contribution to our current understanding of the genotype-phenotype relationship of Wolfram syndrome. These results highlight that alterations in coding sequences result in significant changes in the presentation and severity of Wolfram.

A possible direction to explore these findings would be to study the relationship between variants and Wolfram United Rating Scale (WURS) disease severity scores [22]. Additionally, integrating these results into the current understanding of Wolfram could allow clinicians to explore the possibility of individualized treatments for Wolfram. The implications of this would include improved clinical care for Wolfram patients, with the possibility of missense variants warranting different treatment approaches than frameshift or non-sense variants. It would also allow patients and parents to plan for future outcomes of the disease. Research studies exploring the expressivity of different Wolfram causing gene variants should be undertaken. Due to Wolfram syndrome being caused by numerous different combinations of variants, it has been challenging to aggregate data to determine patterns in expressivity.

Additional directions for research include exploring the correlation between levels of biomarkers and severity of disease. Biomarkers for neurodegeneration, such as neurofilament light chain and myelin basic protein, and inflammatory cytokine levels have been shown to be elevated in patients with Wolfram [23; 24]. The use of functional assays could be helpful in this regard to further explore the role of these biomarkers in the pathogenesis of Wolfram. Establishing a correlation between biomarker levels and phenotype severity will aid in the prediction of clinical outcomes and disease progression in patients with Wolfram syndrome [23].

Although correlations between genotype and phenotype presentations for Wolfram syndrome have been posed in this paper, further research is needed to confirm these conjectures.

## Data Availability

All data produced in the present study are available upon reasonable request to the authors

## Author Contributions

EL, MLDH, and FU conceived the experimental design. NP, EP, SL, LB, PN, WA, CB, SH, MV, VN, MLDH, and FU collected information from patients and databases. EL performed data analysis. MV, EL, and FU wrote the manuscript. All authors edited and reviewed the manuscript.

## Acknowledgments

We thank all the members of the Washington University Wolfram Syndrome Study and Research Clinic for their support (https://wolframsyndrome.wustl.edu) and all the participants in the Wolfram syndrome International Registry and Clinical Study, Research Clinic, and Clinical Trials for their time and efforts.

## Data Availability Statement

The original contributions presented in the study are included in the article/Supplementary Material. Further inquiries can be directed to the corresponding author.

---

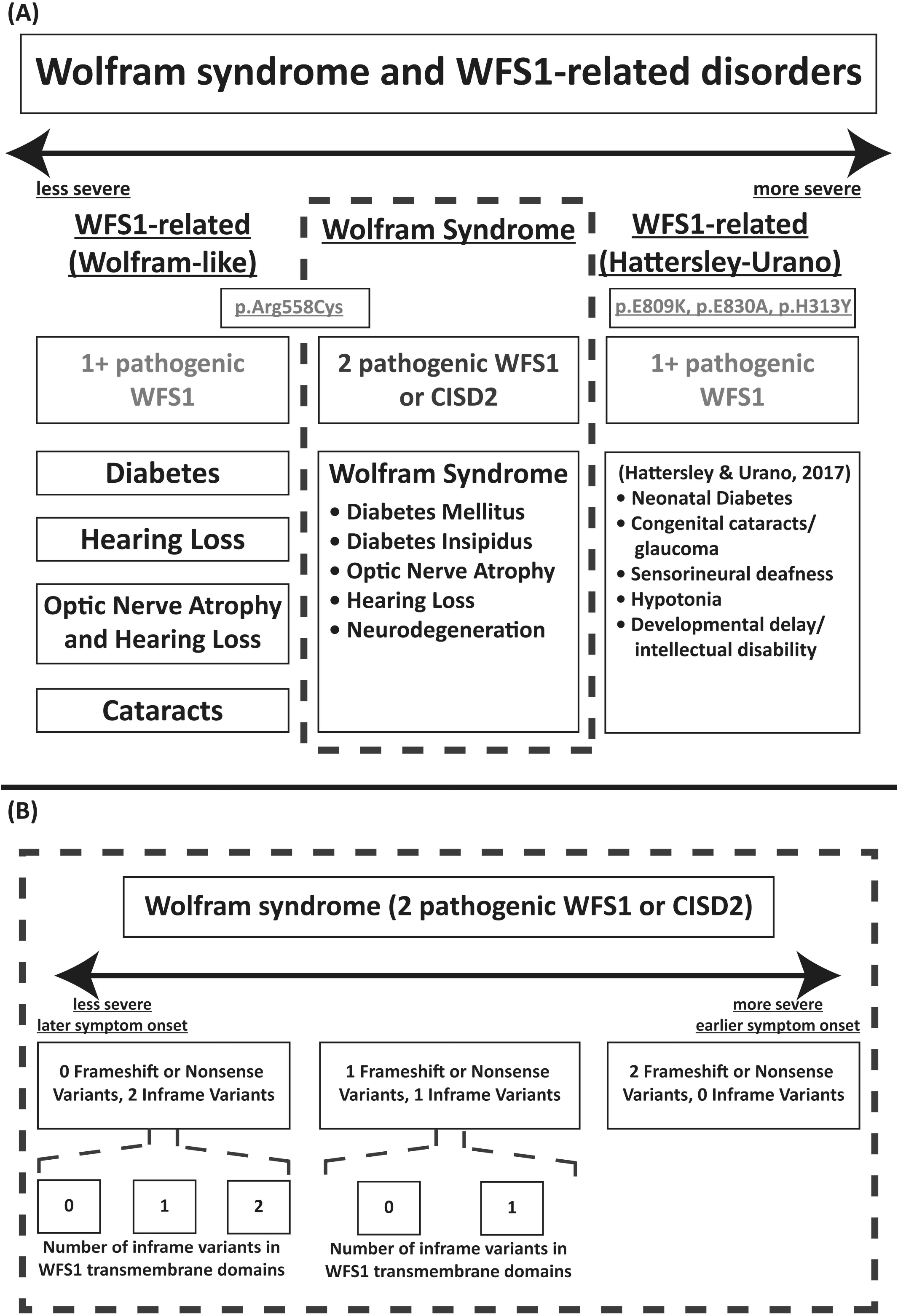

